# The interplay of policy, behavior, and socioeconomic conditions in early COVID-19 epidemiology in Georgia

**DOI:** 10.1101/2021.03.24.21254256

**Authors:** Mallory J. Harris, Ella Tessier-Lavigne, Erin A. Mordecai

## Abstract

To investigate the impact of local public health orders, behavior, and population factors on early epidemic dynamics, we investigated variation among counties in the U.S. state of Georgia. We conducted regressions to identify predictors of (1) local public health orders, (2) mobility as a proxy for behavior, and (3) epidemiological outcomes (i.e., cases and deaths). We used an event study to determine whether social distancing and shelter-in-place orders caused a change in mobility.

Counties at greater risk for large early outbreaks (i.e., larger populations and earlier first cases) were more likely to introduce local public health orders. Social distancing orders gradually reduced mobility by 19% ten days after their introduction, and lower mobility was associated with fewer cases and deaths. Air pollution and population size were predictors of cases and deaths, while larger elderly or Black population were predictors of lower mobility and greater cases, suggesting self-protective behavior in vulnerable populations. Early epidemiological outcomes reflected responses to policy orders and existing health and socioeconomic disparities related to disease vulnerability and ability to socially distance. Teasing apart the impact of behavior changes and population factors is difficult because the epidemic is embedded in a complex social system with multiple potential feedbacks.

## Introduction

In the early stages of an emerging epidemic without existing population immunity or effective vaccines or therapeutics, nonpharmaceutical interventions like non-essential business closures and bans on social gatherings are some of the only effective measures to control disease transmission.^1^ These interventions have been successfully implemented historically and were introduced in many locations at the beginning of the COVID-19 pandemic.^2,3^ Slowing transmission in the early stages of the COVID-19 pandemic has been critical for minimizing deaths and for keeping new hospitalizations below health systems capacity, allowing public health departments to build testing capacity for targeted intervention strategies (i.e., contact tracing), and potentially giving researchers time to develop more effective treatments and vaccines.^4,5^ However, the ability to socially distance is often limited for people with low incomes, including many in communities of color, exacerbating the disproportionate impact of this virus on marginalized groups, which also tend to have higher rates of relevant comorbidities as a result of health inequities and systemic racism (e.g., heightened exposure to air pollution that may worsen outcomes for COVID-19 patients).^6–9^

The first confirmed case of COVID-19 in the United States was reported in late January, 2020.^10^ In the following months, the virus began to spread nationally, often with delayed detection. State level responses varied tremendously, due in part to spatial heterogeneity in virus spread early in the epidemic, as well as differences in perspectives on the virus that often fell along partisan lines.^11,12^ For example, on March 19^th^, 2020, California Governor Newsom introduced the country’s first statewide shelter-in-place order, and all but eight states enacted shelter-in-place orders by April 7^th^.^13^ Most other states followed, and all but eight states enacted shelter-in-place orders by April 7^th^ (Arkansas, Iowa, Nebraska, North Dakota, South Dakota, Oklahoma, Utah, and Wyoming; states that had notably explosive outbreaks months later, in the fall of 2020).^13^ In some cases, when states delayed nonpharmaceutical interventions despite local transmission, county and municipal governments introduced stricter public health orders than those established at the state level.

Georgia presents a case study to understand the local effects of policy at the beginning of the pandemic due to the combination of a relatively early hotspot, delayed statewide action, and a patchwork of earlier local orders. In a national analysis, multiple Georgia counties were identified as particularly vulnerable to COVID-19 due to intersecting socioeconomic and health risk factors.^14^ The first COVID-19 case in Georgia was recorded on March 3^rd^ and by March 27^th^ Albany, Georgia had the third highest per capita death rate of any metro area in the world, following a February superspreading event that was not detected until several weeks later.^10,15^ On March 20^th^, Athens-Clarke County became the first local government in Georgia to issue a shelter-in-place order, while Governor Kemp banned gatherings of more than ten people on March 24^th^ and issued a statewide shelter-in-place on April 3^rd^. ^16–18^ Nineteen additional counties had similar shelter-in-place orders prior to the Governor’s statewide order, while 23 of 159 counties introduced measures to promote social distancing prior to the Governor’s large gathering ban (Fig. 1).^17,19^ Local interventions tended to be clustered in metro-Atlanta counties, but there was some geographic heterogeneity in county-level measures (Fig. 1).

**Figure 1:**
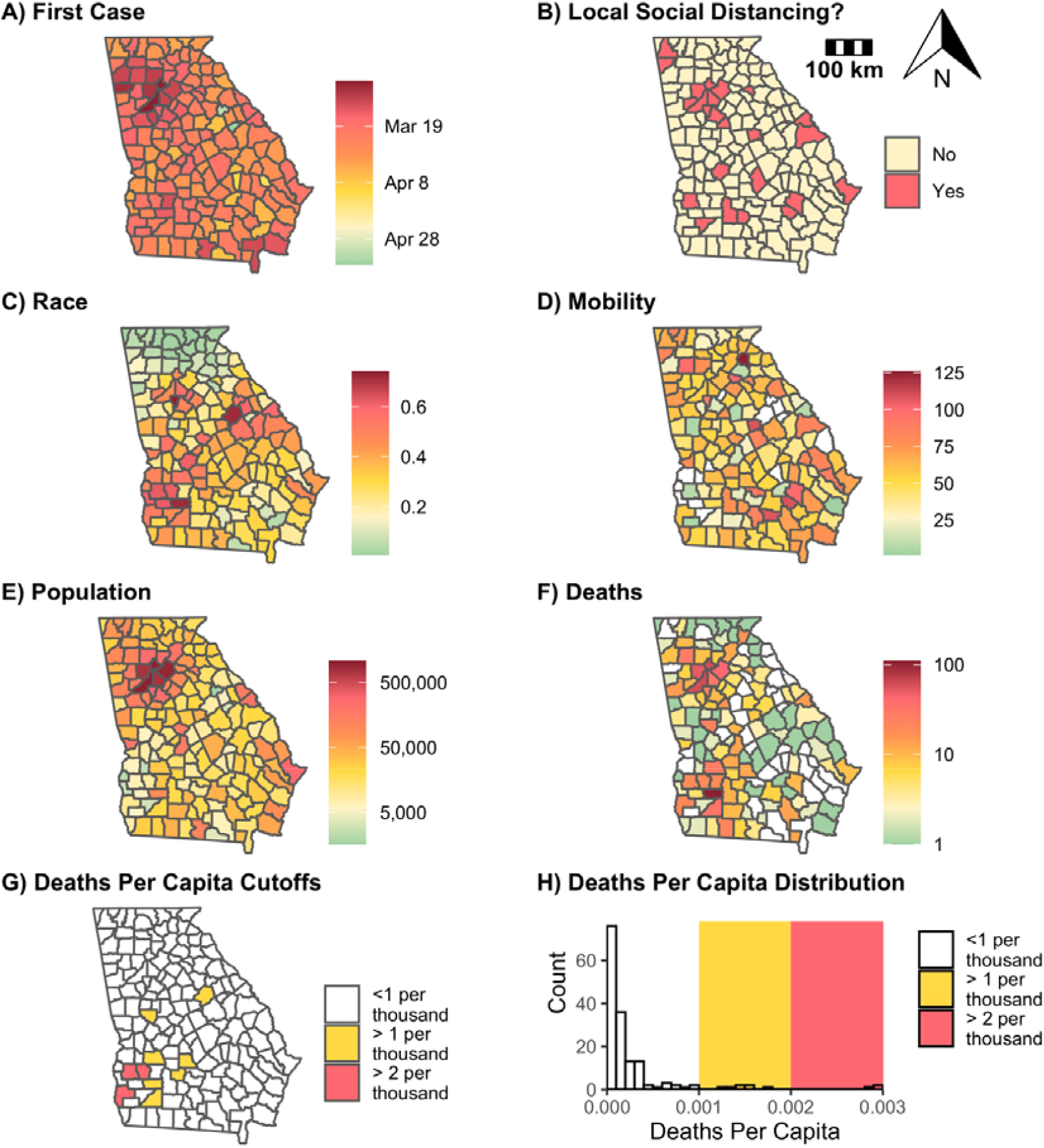
Variation among Georgia counties in first case detection, social distancing orders, race, mobility, population size, and COVID-19 deaths. Counties are shaded according to their values for the given covariate used in regressions: (A) the date the first case was reported, (B) whether a local social distancing order was passed prior to the statewide order, (C) the proportion of the county that identifies as Black, (D) mobility normalized to a pre-pandemic baseline (m50_index), averaged across the final week of the statewide shelter-in-place order, (E) natural log of population size, (F) natural log of cumulative COVID-19 deaths reported in the six weeks following a county’s first case report, and (G) whether per capita COVID-19 deaths exceeded one per thousand (yellow) or two per thousand (pink), which were excluded from regressions in sensitivity analyses. (H) is a histogram that shows the distribution of COVID-19 deaths per capita across Georgia, shaded according to the thresholds for per capita deaths.

It is important to understand the efficacy of county-level ordinances and to identify socioeconomic predictors of worse early outcomes and reduced nonpharmaceutical intervention compliance.^20–22^ These analyses are complicated by the presence of several interrelated covariates that may have bidirectional relationships (e.g., nonpharmaceutical intervention may reduce transmission, but counties may enact these policies because they already have high transmission rates).^20^ For example, counties with low median household income and educational attainment and high unemployment and poverty rates are predicted to have larger working class populations who were limited in their ability to practice nonpharmaceutical interventions, while high housing density and air pollution may also indicate more urbanized areas with more rapid early spread.^21,22^ Identifying predictors of worse early outbreaks and reduced ability to follow nonpharmaceutical interventions could guide future responses to emerging infectious diseases and inform ongoing COVID-19 response strategies.

In this study, we examined the interplay between health and socioeconomic factors, public health orders, mobility as a proxy for behavior, and early COVID-19 epidemic outcomes, some of which may be bidirectional or cyclical, in Georgia at the county level (Fig. 2). Specifically, we asked: (1) What county-level demographic and epidemiological characteristics predict the introduction of local public health orders? (2) Did public health orders decrease mobility? (3) How are socioeconomic factors related mobility? (4) Which socioeconomic, health, and behavioral factors best predict COVID-19 cases and deaths during the early epidemic period? To answer questions one, three, and four, we conducted regressions and used model selection to identify the top predictors of each response variable. To answer the second question, we conducted an event study to quantify the causal impact of public health orders on mobility.

**Figure 2:**
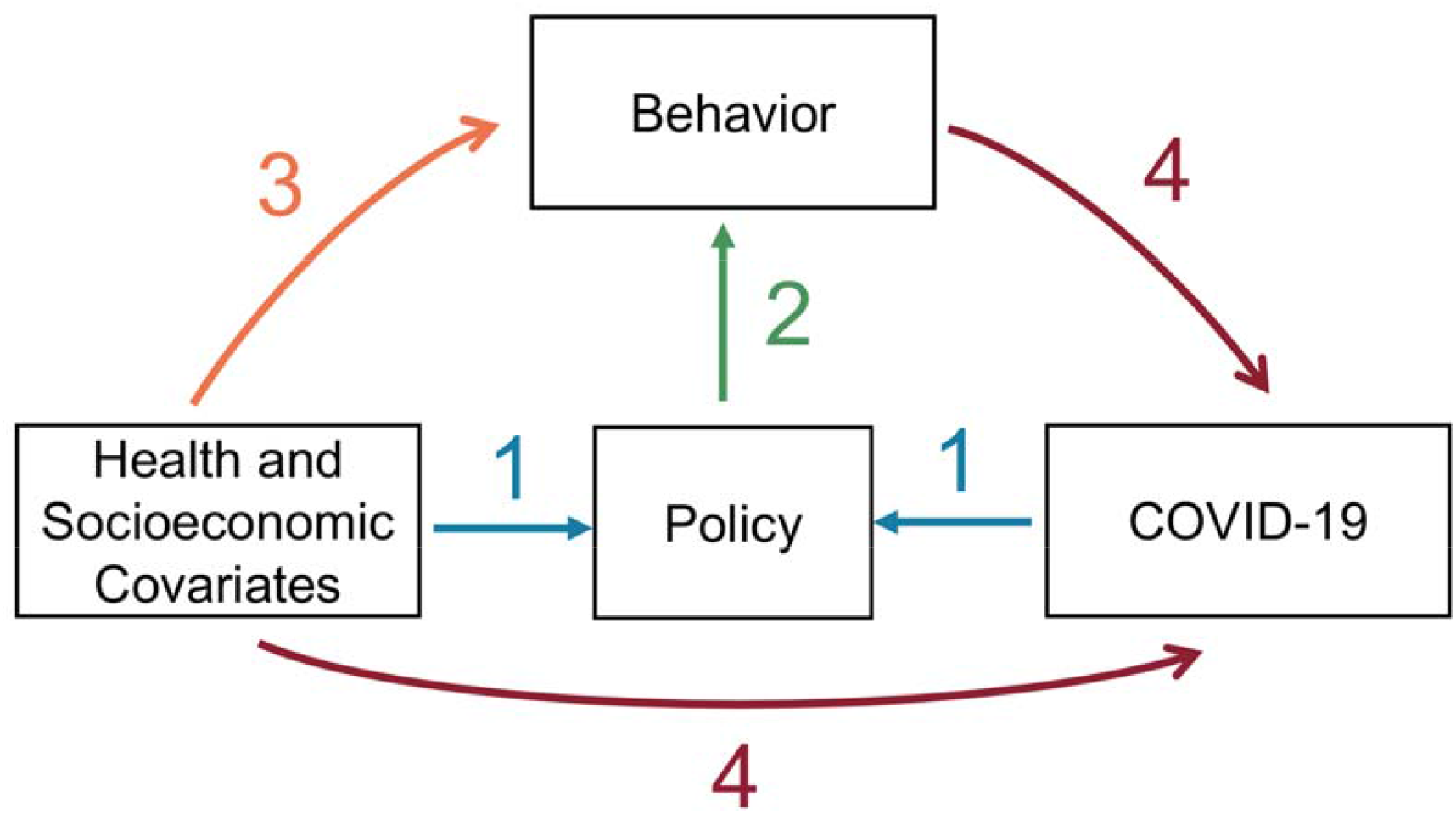
Drivers of COVID-19 epidemiological outcomes—behavior, policy, health, and socioeconomic covariates—are interconnected. Colored arrows correspond with the four-part analyses described here: (1) blue: health and socioeconomic predictors of county-level social distancing or shelter-in-place orders preceding the statewide order (logistic regression), (2) green: effect of social distancing and shelter-in-place policies on mobility as a proxy for behavior (event study), (3) orange: health and socioeconomic predictors of mobility in the final week of April as a proxy for behavior (Gaussian linear regression), (4) red: socioeconomic, health, and mobility predictors of early COVID-19 cases and deaths (negative binomial regression). The color scheme assigned to arrows 1-4 are maintained in the plots pertaining to each of the four components of this study (Fig. 3, Fig. 4, Fig 5., Fig. 6).

## Methods

### Epidemiological Data

We used daily county-level COVID-19 cases and deaths reported by the Georgia Department of Public Health and aggregated in the COVID-19 Data Repository by the Center for Systems Science and Engineering (CSSE) at Johns Hopkins University.^10^ In these analyses, we included cases and deaths reported within four and six weeks, respectively, of each county’s first reported case because we were interested in studying early epidemic outcomes. The additional two weeks for deaths accounts for the lag between case detection and mortality.^23^ We also computed cumulative deaths per capita as of May 21^st^, reflecting transmission prior to the end of the statewide shelter-in-place order.

### Legislative Data

We used daily public health orders implemented at the county based on State Executive Orders, Departments of Education, and other news sources and aggregated in the Center for the Ecology of Infectious Disease at the University of Georgia’s COVID-19-DATA repository.^19^ We defined policies that encourage social distancing in the general population as policies that ban gatherings at non-essential businesses, restrictions on gathering sizes, closures of public use areas, and ordinances that otherwise encourage social distancing. School closures were not included under this definition of social distancing orders as only nine counties implemented local school closures, all within one week of the March 16^th^ statewide school closures, precluding meaningful comparisons. For each county, we defined the beginning of social distancing and shelter-in-place based on the date of the statewide orders if they were enacted prior to any county-level legislation.

### Socioeconomic Data

Population size and the proportion of the county identifying as Black or African American, Hispanic or Latino, Asian, and American Indian and Alaska Native were based on the US Census Bureau’s county-level estimates for 2018.^24^ The proportion of the population identifying as White was excluded from the analysis, as it was highly negatively correlated with the proportion of the population identifying as Black (Supplement: File S1). Population size was log-transformed for all regressions. We also incorporated educational attainment (i.e., proportion of the population with a high school diploma), unemployment, percentage of people below the poverty line, median household income, and housing units per square mile compiled previously from US Census Bureau reports as indicators of socioeconomic status and urbanization.^14^ We calculated county-wide age-weighted infection fatality rate based on age-specific infection fatality rates and the US Census Bureau’s 2018 estimates of the proportion of each county in corresponding age bins.^25,26^

### Partisanship Data

The partisanship of each county was defined as the vote margin in percentage points between the Republican and Democratic candidates for Governor of Georgia in 2018, with more positive values indicating counties with more Republican voters.^27^

### Comorbidity and Health Data

Data on pollution (Particulate Matter PM2.5) and relevant health comorbidities (obesity, coronary heart disease, and diabetes) were compiled previously from the Centers for Disease Control and Environmental Protection Agency.^14^ We collected additional data on asthma from the Georgia Department of Public Health.^28^

### Mobility Data

We computed the proportion of each county’s population that works in another county based on the 2011-2015 American Community Survey Commuting Flows.^29^ To measure variation in mobility over time and space, our metric of behavioral changes related to the pandemic, we used daily county-level statistics based on mobile phone data from Descartes Lab.^30^ The maximum distance traveled from the initial point on each day was recorded for every device and the daily median across devices (m50) in a county was calculated. Normalized daily mobility (*m50*_index) was defined as a proportion of baseline mobility prior to widespread mobility changes in the US.^30^ For regressions, we defined mobility as the mean *m50*_index in the final week of April, corresponding to the end of the shelter in place period. Ten counties were excluded from the analyses because they had no available mobility data (Baker, Calhoun, Clay, Glascock, Hancock, Quitman, Stewart, Taliaferro, Warren, Webster, and Wheeler) (Fig. 1).*Data Analysis*

### Part 1: Predictors of local public health orders: logistic regression

We conducted logistic regression to identify predictors of a county’s having a local social distancing or shelter-in-place order prior to the statewide orders. Covariates were normalized by subtracting the mean and dividing by standard deviation to allow direct comparisons of effect sizes. Forward and backward model selection was conducted to minimize AIC, balancing goodness-of-fit against overfitting.

We tested whether the inclusion of counties with extreme values for COVID-19 deaths per capita skewed our results by performing sensitivity analyses excluding the three counties— involved in an early superspreading event—where per capita death rates exceeded two per thousand (Randolph, Terrell, and Early) or the ten counties where per capita death rates exceeded one per thousand (Randolph, Terrell, Early, Hancock, Turner, Dougherty, Wilcox, Mitchell, Sumter, and Upson) (Fig. 1). We computed Nagelkerke’s pseudo-R^2^ for all models.^31,32^ All analyses were conducted in R statistical software version 4.0.0.

### Part 2: Effect of public health orders on mobility: event study

We used an event study framework to understand the effect of public health orders (social distancing or shelter-in-place) on mobility at the county level. This approach seeks to identify changes in time series data following a pre-specified event. For event study analyses, we included the ten days prior to and following the legislation’s introduction in each county, spanning the time difference between the statewide social distancing and shelter-in-place orders to isolate the effects of the two orders. The covariate NPI_day was defined as follows:

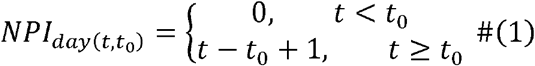

where *t* is the time in days and *t*_0_ is the date a particular order was introduced.

We used a fixed effect model to adjust for variation due to county and date and to quantify both the binary effects of nonpharmaceutical interventions and the effect of days since a nonpharmaceutical interventions was introduced. The model formulation was:

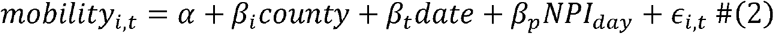

Where *α* is an intercept, *β*’s are coefficients for corresponding covariates, and *ϵ* is an error term for each county *i* and date *t*. In addition to the ten counties excluded from regression due to no mobility data, seven more counties were excluded from the both event studies due to incomplete mobility data for the study period (Chattahoochee, Marion, Randolph, Schley, and Twiggs) and Montgomery county was excluded only from the event study for shelter-in-place orders.

### Part 3: Predictors of mobility: Gaussian linear regression

We examined the relationship between socioeconomic variables and average mobility in the last week of April using a Gaussian linear regression to identify predictors of nonpharmaceutical intervention compliance. Model selection was conducted as described in part one.

### Part 4: Predictors of early epidemiological outcomes: negative binomial regression

We identified the primary socioeconomic, health, and behavioral predictors of early epidemic outcomes by fitting negative binomial regressions to reported COVID-19 cases and death. Both responses were count variables that were overdispersed relative to the expected variance in a Poisson distribution. We performed model selection and computed Nagelkerke’s pseudo-R^2^ as described in part one.^31,32^

## Results

### Part 1: Predictors of local public health orders: logistic regression

Counties with a greater share of Democratic voters and larger populations were more likely to introduce social distancing orders (Supplement: Table S1, Fig. 3). Counties with earlier first case detection and larger populations were more likely to pass shelter-in-place orders. Most of these counties contain large municipalities (e.g., Atlanta, Athens, and Macon). All findings were robust to the inclusion or exclusion of counties with high per capita deaths (>1 or 2 per 1000). Socioeconomic and demographic variables captured less variation in the passage of local social distancing orders (pseudo-R^2^: 0.25-0.31) compared to local shelter-in-place orders (pseudo-R^2^: 0.51-0.54) (Supplement: Table S1).

**Figure 3:**
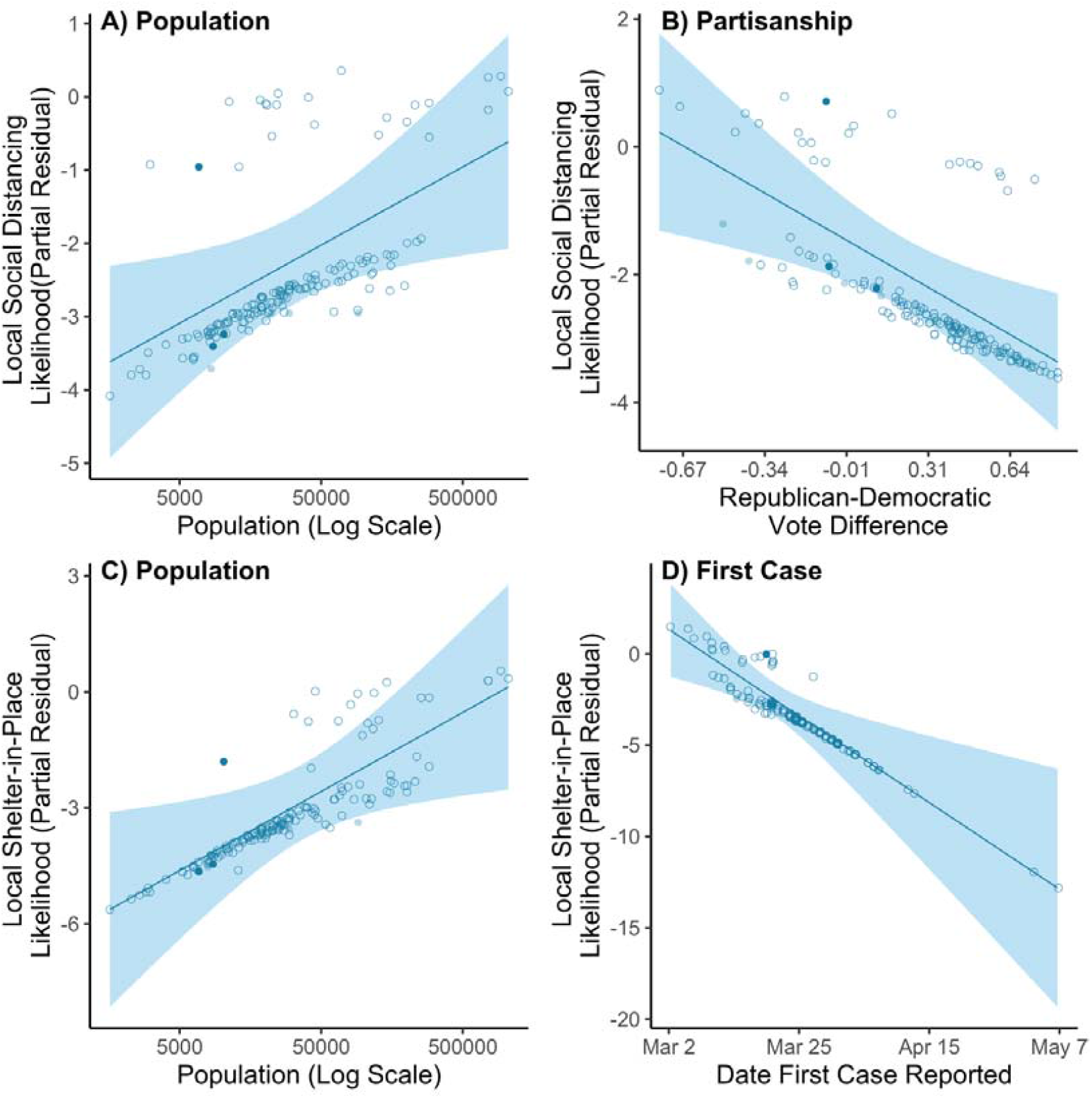
Counties with larger populations were more likely to enact local social distancing and shelter-in-place orders. Partial residual plots with lines giving the estimated relationship between predictors and relative likelihood of a local nonpharmaceutical intervention order, with the 95% confidence interval is indicated as a shaded band. The points indicate the marginal relationship at the county level between predictors and relative logit-transformed likelihood of a local nonpharmaceutical intervention, after adjusting for all other predictors selected in the best fit model. The top row shows the two most significant predictors of a local social distancing order: logged population size (Population) and percent point difference of Republican and Democratic vote share in 2018 gubernatorial election, with more negative values indicating a higher proportion of Democratic voters (Partisanship). The bottom row shows the two most significant predictors of a local shelter-in-place order: logged population size (Population) and date first case in county was detected (First Case). Open circles indicate counties with less than one death per thousand people, while light and dark shaded circles indicate counties with outlying values for per capita deaths (thresholds of one or two deaths per thousand people, respectively).

### Part 2: Effect of public health orders on mobility

Mobility decreased by 19% (P<0.001) in the ten days following the introduction of a social distancing order (Supplement: Table S2). We observed 21 instances (county-days) where mobility exceeded the county- and date-adjusted mean for the event study period by over 35%—which we designated as mobility extremes—and all occurred prior to the introduction of local social distancing orders (Fig. 4).

**Figure 4:**
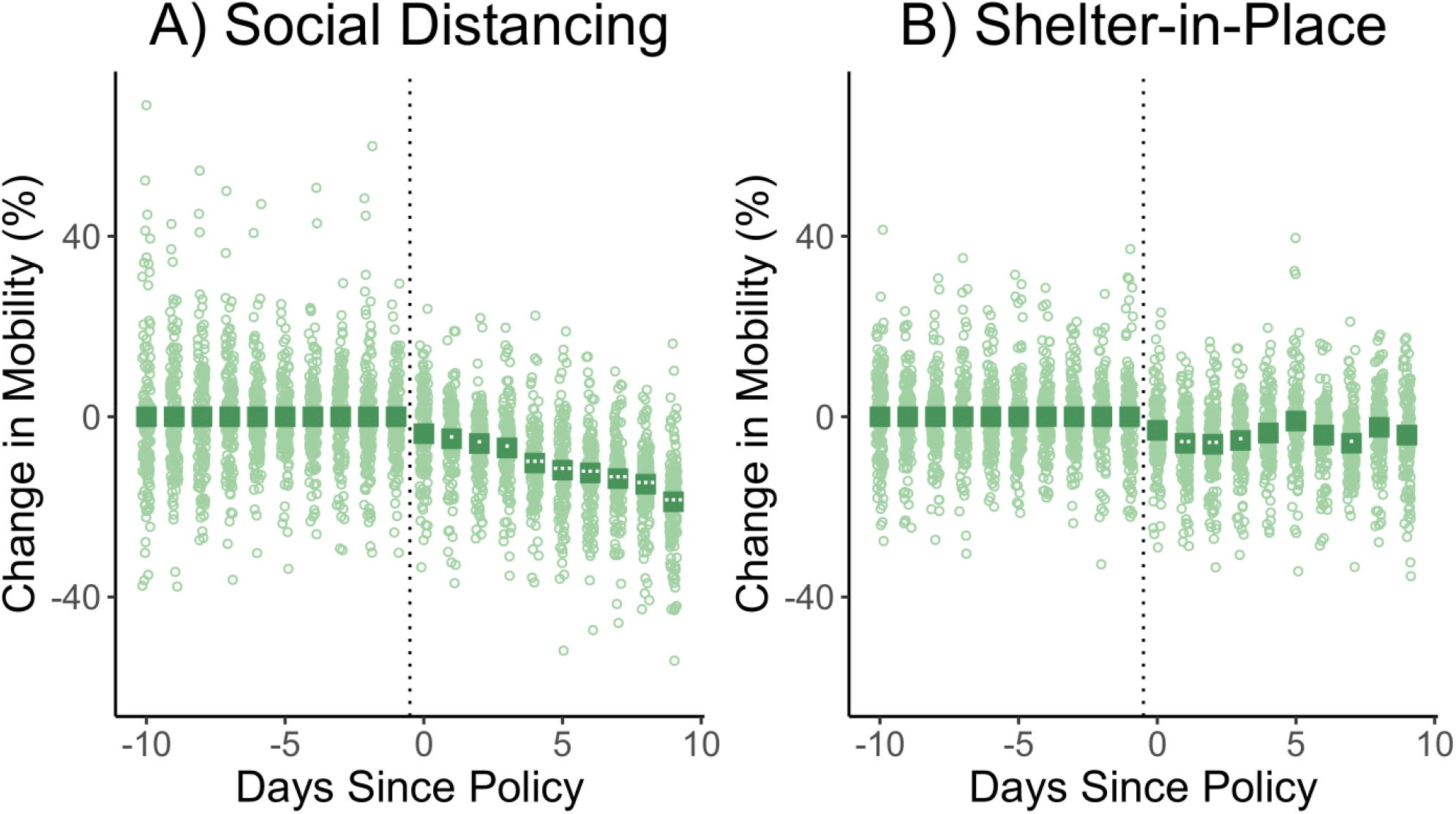
Social distancing orders gradually reduced mobility by up to 19%, while shelter-in-place orders had only a short-term marginal effect for days 2-4. The coefficients of the event studies for social distancing policies (left) and shelter-in-place orders (right) are given as squares across the ten days preceding and following the introduction of the policy, with the day the order was introduced indicated with a vertical dotted line. The significance of the coefficients is indicated by the number of white dots within each square (⍰: *P*<0.05; ⍰⍰: *P* <0.01, ⍰⍰⍰: *P* <0.001). The green circles indicate the marginal effect of policy on mobility by date in each county, after adjusting for county and date fixed effects.

All counties had social distancing orders prior to shelter-in-place orders; statewide social distancing orders were introduced ten days before the statewide shelter-in-place order. Therefore, the event study involving shelter-in-place orders indicates the marginal effect of shelter-in-place orders after accounting for social distancing orders. Overall, although mobility was significantly reduced two to five days after shelter-in-place orders were passed, we did not detect a sustained marginal effect of shelter-in-place orders on mobility, after accounting for the effects of social distancing orders already in place (Supplement: Table S2, Fig. 4). County and date fixed effects are reported in Tables S3-4.

### Part 3: Socioeconomic predictors of mobility: Gaussian linear regression

Age, income, and the proportion of the population identifying as Black were all significant negative predictors of mobility (Supplement: Table S5, Fig. 5). Mobility declined by 20 percentage points for every 0.0052 increase in age-weighted infection fatality rate, $5,207 increase in median household income, or 39 percentage point increase in the Black proportion of the population. There was little variation in effect sizes when counties with outlying per capita death rates were excluded. Of the ten counties where per capita deaths exceeded one per thousand, all had median household income below the statewide mean ($44,000), nine had Black population proportions above the statewide average of 0.30 (and six were majority Black), and seven had age-weighted infection fatality rates above the statewide average of 0.011 (Fig. 5). The model only captured 11-13% of observed variation in mobility (Supplement: Table S5).

**Figure 5:**
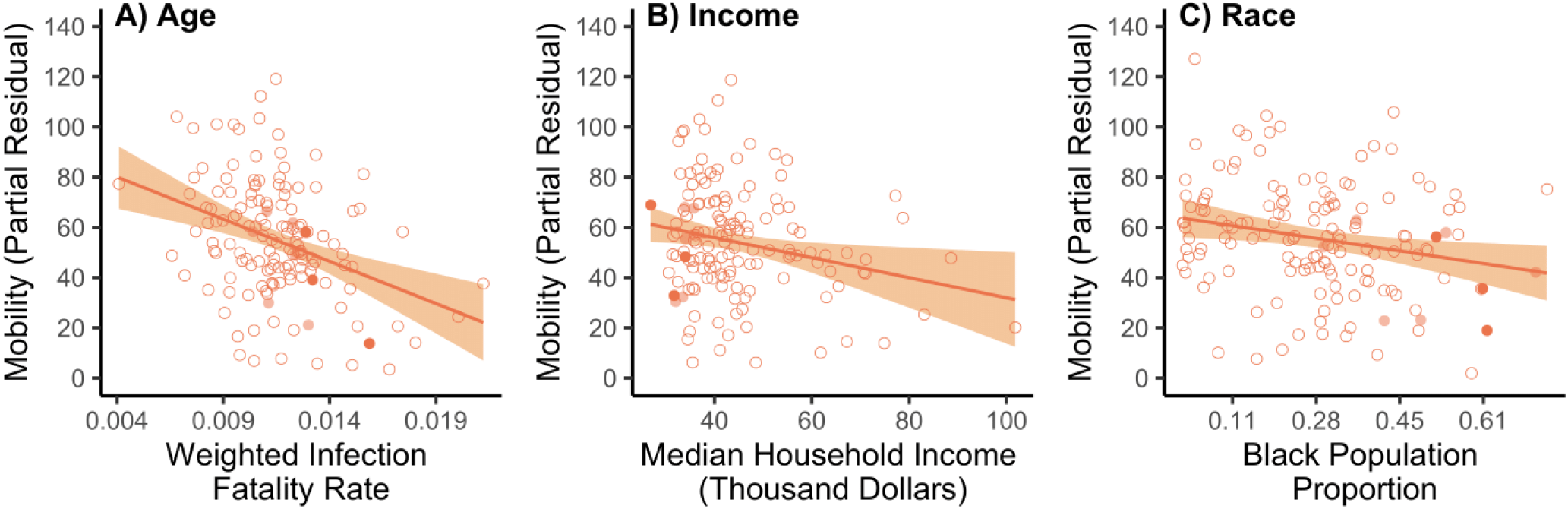
Higher age, median household income, and Black proportion of population all corresponded to lower mobility in the final week of the statewide shelter-in-place order. Partial residual plots with lines indicating the estimated relationship between predictors and mean mobility in the final week of the statewide shelter-in-place order, while the 95% confidence interval is indicated as a shaded band. The points indicate the marginal relationship at the county level between predictors and mobility, after adjusting for all other predictors selected in the best fit model. All predictors selected in the best fit model are displayed: age-weighted infection fatality rates (Age), median household income (Income), and percent of the population that is Black (Race). Open circles indicate counties with less than one death per thousand people, while light and dark shaded circles indicate counties with outlying values for per capita deaths (thresholds of one or two deaths per thousand people, respectively).

### Part 4: Socioeconomic, health, and mobility predictors of early epidemiological outcomes: negative binomial regression

Counties with larger populations and higher air pollution had significantly more cases and deaths across all models, while greater mobility was a significant positive predictor of cases and deaths only in the models that excluded the ten counties where per capita deaths exceed one per thousand (Supplement: Table S6, Fig. 6). Counties with greater proportions of the population who were elderly or living below the poverty line and lower rates of coronary heart disease reported more cases, while counties with lower educational attainment and earlier first cases reported more deaths. Additional health and socioeconomic covariates were included in some of the models selected by AIC, but their effect sizes were not significantly different from zero unless counties with per capita deaths greater than one per thousand were excluded (e.g., diabetes, asthma). All predictors included in the models explained 67-73% of the variation in cases and 49-53% of the variation in deaths, where ranges depended on the subset of outlier counties that were included.

**Figure 6:**
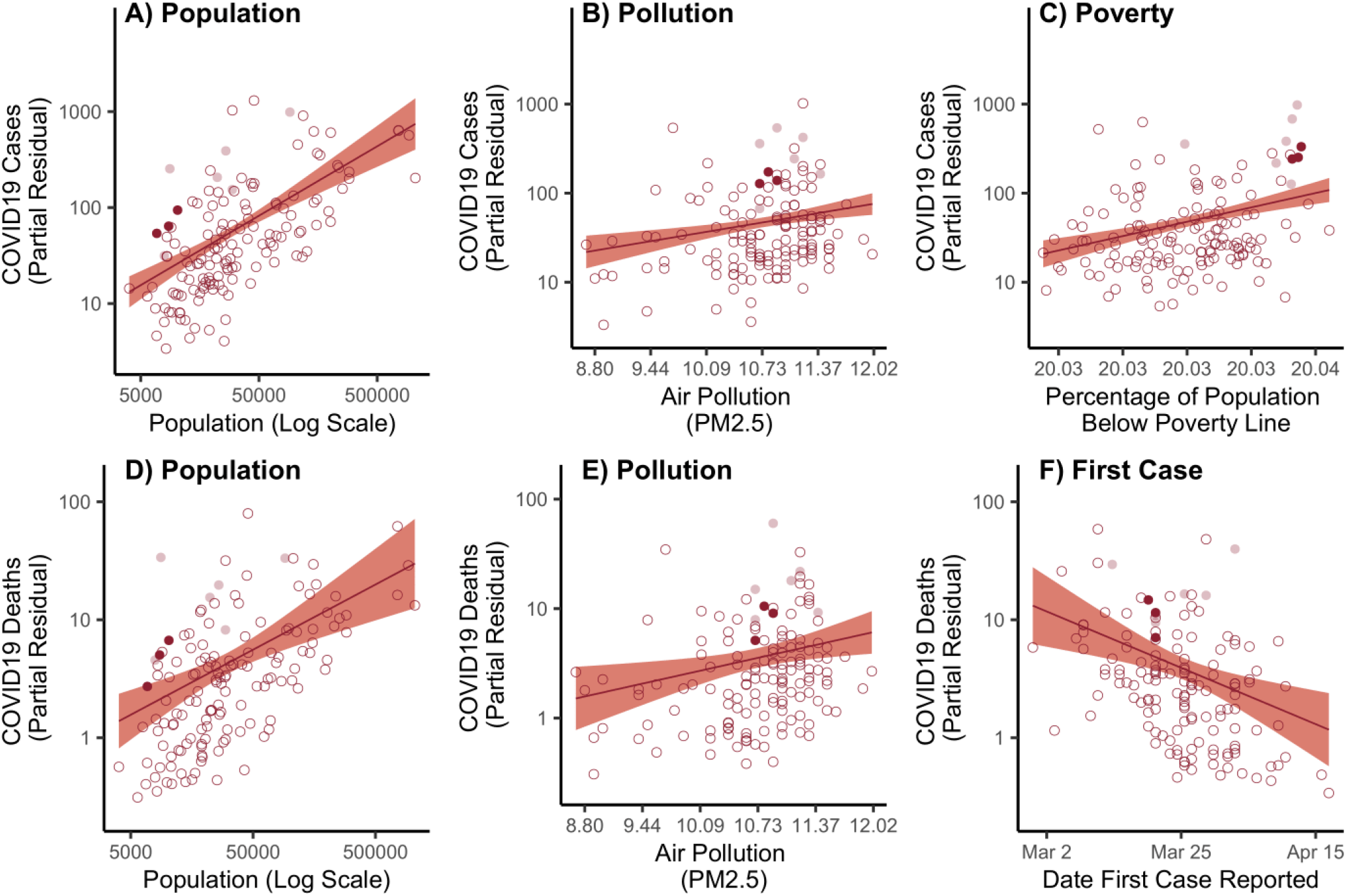
Larger population size, higher pollution, and higher poverty rates corresponded to more COVID-19 cases at the county level. Larger population size, higher pollution, and earlier first case reported corresponded to more COVID-19 deaths at the county level. Partial residual plots with lines giving the estimated multiplicative relationship between predictors and COVID-19 cases and deaths, with the 95% confidence interval is indicated as a shaded band. The points indicate the marginal multiplicative relationship at the county level between predictors and COVID-19 cases (top) or deaths (bottom), after adjusting for all other predictors selected in the best fit model. The top row shows the three most significant predictors of early COVID-19 cases: logged population size (Population), annual average ambient PM2.5 concentration (Pollution), and percentage of population living below poverty line (Poverty). The bottom row shows the three most significant predictors of early COVID-19 deaths: logged population size (Population), annual average ambient PM2.5 concentration (Pollution), and date first case in county was detected (First Case). Open circles indicate counties with less than one death per thousand people, while light and dark shaded circles indicate counties with outlying values for per capita deaths (thresholds of one or two deaths per thousand people, respectively).

## Discussion

Social distancing orders successfully reduced mobility, and lower mobility was associated with fewer COVID-19 deaths in most Georgia counties. Mobility gradually declined by 19% (95% CI: - 27% 10%) over ten days after social distancing orders were introduced, suggesting that, with some lag, these orders contributed to behavioral changes that may be indicative of social distancing (Supplement: Table S5, Fig. 4). Conversely, we found that undoing this level of mobility change—i.e., a 19% increase during the final week of shelter-in-place—would be associated with a 17% (95% CI: 1-35%) increase in COVID-19 deaths or 10% (95% CI 0-20%) increase in cases in the counties where per capita deaths were fewer than one per thousand (Supplement: Table S6, Fig. 6). Moreover, we did not detect a significant marginal effect on mobility from shelter-in-place orders after social distancing orders had already been introduced.

Mobility data and the analyses presented here may not fully capture behavioral changes linked to nonpharmaceutical interventions. For example, while mobility did not significantly decrease following shelter-in-place orders when social distancing orders were already in place, Georgians may have reduced social contacts within a small radius of their homes following the shelter-in-place order. On the other hand, the calculated reduction in mobility following social distancing orders may not be directly proportional to the reduction in social contacts and high-risk transmission settings (including indoor gatherings without face masks). This analysis does not capture the effects of additional public health measures (e.g., mask mandates and school closures) or behavioral changes prior to the public health orders. This approach to understanding effects of nonpharmaceutical interventions does not capture spillover effects from geographically and socially connected counties, which could expand or distort the influence of local public health orders.^33^ However, epidemiological models fit to cases, deaths, and mobility data similar to those used here have demonstrated that time-varying transmission rates can be captured accurately using mobility data.^34^

In addition to the association with mobility, epidemiological outcomes were predicted by demographic, socioeconomic, and health factors. As expected, counties with larger populations sustained larger outbreaks because the rate of new infections is directly proportional to the number of susceptible people. Higher air pollution was also associated with more cases and deaths, potentially due to more rapid spread in more urbanized counties and/or to worse outcomes in communities with higher rates of health conditions linked to air pollution exposure.^9^ However, contrary to our expectation, we found that the prevalence of comorbidities that are known to worsen individual outcomes for patients with COVID-19 (e.g., obesity and asthma) were not significant predictors of deaths or were unexpectedly negatively associated with early cases and deaths (e.g., coronary heart disease), potentially because they are confounded with factors like income and race. Counties with a larger share of residents who were Black or living below the poverty line experienced more cases and/or deaths, a pattern that may reflect disparities and systemic injustices connected to racism in healthcare, housing, and occupation in Georgia and across the United States.^6,8,21,35^ These covariates may also indicate counties that have larger populations of workers who are unable to work from home and lack sufficient workplace protections.^12,36,37^ Counties with lower median household income had higher mobility, potentially supporting this hypothesis (Supplement: Table S5, Fig. 5). While the proportion of the population identifying as either Hispanic or Latino, Asian, or American Indian and Alaska Native was not a significant predictor of cases, deaths, or mobility at the population level, more data and detailed studies are necessary to understand the impacts of discrimination and injustice across different ethnic and racial groups (Supplement: File S1). Identifying the mechanisms and relative importance of these potential drivers of disparate outcomes is critical for addressing the disproportionate impact of COVID-19 on marginalized communities. Notably, almost all counties with especially high outlying values for per capita deaths at the beginning of the epidemic had median household incomes below and Black population shares above the statewide averages (Fig. 3).

Georgia’s statewide response was relatively delayed compared to most U.S. states and supplemented with more restrictive local responses, which allowed us to compare the effects of various public health orders across counties. However, we found support for the hypothesis that the relationship between nonpharmaceutical interventions and early epidemiological outcomes was bidirectional, a trend that has also been observed in counties that mandated wearing face coverings later in the epidemic.^20^ Counties with earlier detection of cases and larger populations (predictive of larger outbreaks) tended to pass local orders before the statewide order (Supplement: Table S1, Fig. 3), and these orders led to reduced mobility, a predictor of fewer cases (Supplement: Table S2, Supplement: Table S6, Fig. 4, Fig. 6). At the county level, having a higher proportion of Black or elderly residents was predictive of both lower mobility and more cases, suggesting self-protective behavior in vulnerable groups and a tendency early in the pandemic to detect more severe cases in populations with higher rates of health comorbidities (Supplement: Table S5, Supplement: Table S6, Fig. 5, Fig. 6). Separating the causes and effects of differences in social distancing orders, mobility, and transmission using techniques such as instrumental variables will be important in assessing the efficacy of nonpharmaceutical intervention orders.

This analysis could be extended to more locations, and Georgia’s heterogeneous response could be compared to states like California, which had an early statewide shelter-in-place order. Focusing this analysis within a single state at the beginning of the pandemic allows us to quantify initial spread and to assess the efficacy of interventions related to reducing contacts, in addition to understanding risk factors for large outbreaks at a time when treatments and control measures were especially limited. However, testing limitations and lack of early knowledge about the virus may have contributed to substantial underreporting of cases, especially in rural counties lacking public health infrastructure. As statewide orders were lifted, county governments in Georgia and across the country became increasingly responsible for containing local outbreaks.^38–40^ Local governments will therefore need to understand the impact of these orders and identify county-level features that may affect outbreak risk and nonpharmaceutical intervention implementation to respond to this ongoing pandemic and other emerging infectious diseases.

Infectious disease outbreaks are intimately dependent on the societal settings in which they occur, and COVID-19 is no exception. Demographics, health, economic resources, and social power—and disparities in these factors—within communities affect both their vulnerability to and responses to disease outbreaks. Here, we showed that while social distancing orders did reduce mobility (Supplement: Table S2, Fig. 4), and reduced mobility in turn reduced COVID-19 cases in most counties (Supplement: Table S6, Fig. 6), the efficacy of these nonpharmaceutical interventions was mediated by the will of municipal and state governments to impose, and ability of community members to observe, public health orders. Specifically, counties with earlier epidemics, larger populations, and greater shares of Democratic voters were more likely to introduce social distancing or shelter-in-place orders (Supplement: Table S1, Fig. 3), while mobility was lowest in counties with older residents, higher income, and larger Black populations (Supplement: Table S5, Fig. 5). While changing mobility almost certainly affected COVID-19 transmission, this was one of many factors associated with epidemiological outcomes (Supplement: Table S6, Fig. 6). Because these factors are interconnected through both causal linkages and correlations driven by underlying societal structures and inequities (Fig. 2), it is impossible to completely disentangle the causal effects from observational data. However, this work illustrates the imperative need to consider interconnected policy, behavioral responses, socioeconomic, and demographic conditions in designing and evaluating policy to combat emerging epidemics.

## Data Availability

Data and code are available on Github.

https://github.com/mjharris95/GA-COVID

## Data Accessibility

Data and code are available on Github at https://github.com/mjharris95/GA-COVID

## Acknowledgments

The authors thank Noah Rosenberg for his insightful comments. EAM was supported by the National Science Foundation (DEB-1518681 and DEB-2011147, with support from the Fogarty International Center), the National Institute of General Medical Sciences (R35GM133439), and the Terman Award. ETL was support by the Stanford King Center for Global Development. MJH was supported by the Knight-Hennessy Scholars Program

## Declaration of interests

The authors declare no competing interests.

## Notes

### Competing Interest Statement

The authors have declared no competing interest.

### Summary of Updates

Update x-axis for figure 6C

## References

1. World Health Organization Writing Group, Bell D, Nicoll A, et al. Non-pharmaceutical interventions for pandemic influenza, national and community measures. Emerg Infect Dis. 2006;12(1):88–94. doi:10.3201/eid1201.051371

2. Hatchett RJ, Mecher CE, Lipsitch M. Public health interventions and epidemic intensity during the 1918 influenza pandemic. Proc Natl Acad Sci. 2007;104(18):7582. doi:10.1073/pnas.0610941104

3. Pan A, Liu L, Wang C, et al. Association of public health interventions with the epidemiology of the COVID-19 outbreak in Wuhan, China. JAMA. 2020;323(19):1915–1923. doi:10.1001/jama.2020.6130

4. Davies NG, Kucharski AJ, Eggo RM, et al. Effects of non-pharmaceutical interventions on COVID-19 cases, deaths, and demand for hospital services in the UK: a modelling study. Lancet Public Health. 2020;5(7):e375–e385. doi:10.1016/S2468-2667(20)30133-X

5. Tuite AR, Fisman DN, Greer AL. Mathematical modelling of COVID-19 transmission and mitigation strategies in the population of Ontario, Canada. CMAJ. 2020;192(19):E497–E505. doi:10.1503/cmaj.200476

6. Gray DM, Anyane-Yeboa A, Balzora S, Issaka RB, May FP. COVID-19 and the other pandemic: populations made vulnerable by systemic inequity. Nat Rev Gastroenterol Hepatol. Published online June 15, 2020:1-3. doi:10.1038/s41575-020-0330-8

7. Maroko AR, Nash D, Pavilonis BT. COVID-19 and inequity: A comparative spatial analysis of New York City and Chicago hot spots. J Urban Health. 2020;97(4):461.

8. Williams DR, Cooper LA. COVID-19 and health equity—a new kind of “herd immunity.” JAMA. 2020;323(24):2478–2480. doi:10.1001/jama.2020.8051

9. Wu X, Nethery RC, Sabath MB, Braun D, Dominici F. Air pollution and COVID-19 mortality in the United States: Strengths and limitations of an ecological regression analysis. Sci Adv. 2020;6(45):eabd4049. doi:10.1126/sciadv.abd4049

10. Johns Hopkins University Center for Systems Science and Engineering. 2019 Novel Coronavirus COVID-19 (2019-NCoV) Data Repository.; 2020. Accessed July 14, 2020. https://github.com/CSSEGISandData/COVID-19

11. Allcott H, Boxell L, Conway J, Gentzkow M, Thaler M, Yang D. Polarization and public health: Partisan differences in social distancing during the coronavirus pandemic. J Public Econ. 2020;191:104254. doi:10.1016/j.jpubeco.2020.104254

12. Christensen SR, Pilling EB, Eyring JB, Dickerson G, Sloan CD, Magnusson BM. Political and personal reactions to COVID-19 during initial weeks of social distancing in the United States. PLOS ONE. 2020;15(9):e0239693. doi:10.1371/journal.pone.0239693

13. Courtemanche C, Garuccio J, Le A, Pinkston J, Yelowitz A. Strong social distancing measures in the United States reduced the COVID-19 growth rate. Health Aff (Millwood). Published online 2020:1237-1246.

14. Chin T, Kahn R, Li R, et al. US-county level variation in intersecting individual, household and community characteristics relevant to COVID-19 and planning an equitable response: A cross-sectional analysis. BMJ Open. 2020;10(9):e039886. doi:10.1136/bmjopen-2020-039886

15. Cohn N, Katz J, Sanger-Katz M, Quealy K. Four Ways to Measure Coronavirus Outbreaks in U.S. Metro Areas. The New York Times. https://www.nytimes.com/interactive/2020/03/27/upshot/coronavirus-new-york-comparison.html. Published March 27, 2020. Accessed February 25, 2021.

16. Girtz K. An Ordinance for the Second Declaration of a Local State of Emergency Related to COVID-19; And for Other Purposes.; 2020. https://www.accgov.com/DocumentCenter/View/67293/March-20-COVID-19-Ordinance?bidId=

17. Kemp B.; 2020. Accessed July 7, 2020. https://gov.georgia.gov/document/2020-executive-order/03232001/download

18. Kemp B. Executive Order to Ensure a Safe and Healthy Georgia.; 2020. Accessed July 7, 2020. https://gov.georgia.gov/document/2020-executive-order/04022001/download

19. Evans M, Richards R, Willoughby A, et al. CEIDatUGA/COVID-19-DATA.; 2020. Accessed July 6, 2020. https://github.com/CEIDatUGA/COVID-19-DATA

20. Dyke MEV. Trends in county-level COVID-19 incidence in counties with and without a mask mandate — Kansas, June 1–August 23, 2020. MMWR Morb Mortal Wkly Rep. 2020;69:1777–1781.

21. van Holm EJ, Wyczalkowski CK, Dantzler PA. Neighborhood conditions and the initial outbreak of COVID-19: the case of Louisiana. J Public Health. Published online 2020:1-6. doi:10.1093/pubmed/fdaa147

22. Jay J, Bor J, Nsoesie EO, et al. Neighbourhood income and physical distancing during the COVID-19 pandemic in the United States. Nat Hum Behav. 2020;4(12):1294–1302. doi:10.1038/s41562-020-00998-2

23. Gaythorpe K, Imai N, Cuomo-Dannenburg G, et al. Report 8: Symptom Progression of COVID-19. Imperial College London; 2020. doi:10.25561/77344

24. Race: Annual county resident population estimates by age, sex, race, and hispanic origin: April 1, 2010 to July 1, 2018 (CC-EST2018-ALLDATA). Published online June 2020. https://www2.census.gov/programs-surveys/popest/datasets/2010-2019/counties/totals/

25. Verity R, Okell LC, Dorigatti I, et al. Estimates of the severity of coronavirus disease 2019: a model-based analysis. Lancet Infect Dis. 2020;20(6):669–677. doi:10.1016/S1473-3099(20)30243-7

26. Annual county and resident population estimates by selected age groups and sex: April 1, 2010 to July 1, 2018 (CC-EST2018-AGESEX). Published online June 2020. https://www2.census.gov/programs-surveys/popest/datasets/2010-2018/counties/totals/co-est2018-alldata.csv

27. Crittenden RA. November 6, 2018 general election Official results. Published online November 17, 2018. https://results.enr.clarityelections.com/GA/91639/222278/reports/detailxls.zip

28. Cheng V, Clarkson L, Lopez F, Chambers S. Georgia asthma surveillance report. Published online April 2012. Accessed July 8, 2020. https://dph.georgia.gov/sites/dph.georgia.gov/files/related_files/site_page/2012%20Asthma%20Burden%20Report.pdf

29. 2011-2015 5-year ACS commuting flows. Published online 2015. https://www.census.gov/data/tables/2015/demo/metro-micro/commuting-flows-2015.html

30. Warren M, Skillman S. Mobility changes in response to COVID-19. Published online March 2020. arxiv.org/abs/2003.14228

31. Magee L. R^2 measures based on Wald and likelihood ratio joint significance tests. Am Stat. 1990;44:250–253. doi:10.1080/00031305.1990.10475731

32. Dabao Z. Rsq: R-Squared and Related Measures.; 2020.

33. Holtz D, Zhao M, Benzell SG, et al. Interdependence and the cost of uncoordinated responses to COVID-19. Proc Natl Acad Sci. 2020;117(33):19837–19843. doi:10.1073/pnas.2009522117

34. Kain MP, Childs ML, Becker AD, Mordecai EA. Chopping the tail: how preventing superspreading can help to maintain COVID-19 control. medRxiv. Published online July 3, 2020:2020.06.30.20143115. doi:10.1101/2020.06.30.20143115

35. Azar KMJ, Shen Z, Romanelli RJ, et al. Disparities in outcomes among COVID-19 patients in a large health care system in California. Health Aff (Millwood). 2020;39(7):1253–1262. doi:10.1377/hlthaff.2020.00598

36. Yancy CW. COVID-19 and African Americans. JAMA. 2020;323(19):1891–1892. doi:10.1001/jama.2020.6548

37. Czeisler MÉ, Tynan MA, Howard ME, et al. Public Attitudes, Behaviors, and Beliefs Related to COVID-19, Stay-at-Home Orders, Nonessential Business Closures, and Public Health Guidance — United States, New York City, and Los Angeles, May 5–12, 2020. Morb Mortal Wkly Rep. 2020;69(24):751–758. doi:10.15585/mmwr.mm6924e1

38. Johnson V. Emergency Order Requiring That Face Coverings or Masks Be Worn in Public in the City of Savannah During the COVID-19 Public Health Emergency.; 2020. Accessed July 10, 2020. https://www.savannahga.gov/DocumentCenter/View/19649/June-30-signed-emergency-order?bidId=

39. Lance Bottoms K. Executive Order Number 2020-113.; 2020. https://www.atlantaga.gov/Home/ShowDocument?id=47225

40. California Department of Public Health. County Data Monitoring - Step 2. Accessed July 10, 2020. https://www.cdph.ca.gov/Programs/CID/DCDC/Pages/COVID-19/CountyMonitoringDataStep2.aspx

