## Supplement for "The interplay of policy, behavior, and socioeconomic conditions in early COVID-19 epidemiology in Georgia"

**File S1: Evidence of correlations between racial and ethnic covariates to justify covariate selection.**

In order to determine which racial and ethnic covariates to include in our models, we first identified highly correlated covariates. The proportions of the population identifying as White or Black in a given county were strongly correlated (r = -0.99). We choose to focus on the proportion of Black people in the population based on evidence of increased risk for COVID-19 infection and mortality resulting from health and economic disparities connected to racial discrimination,^1–4^ The remaining three covariates (proportion of the population identifying as Asian, Hispanic or Latino, or American Indian and Alaska Native) are not included in the best fitting models following model selection, meaning that they were not significant predictors of cases, deaths, or mobility at the county-level.

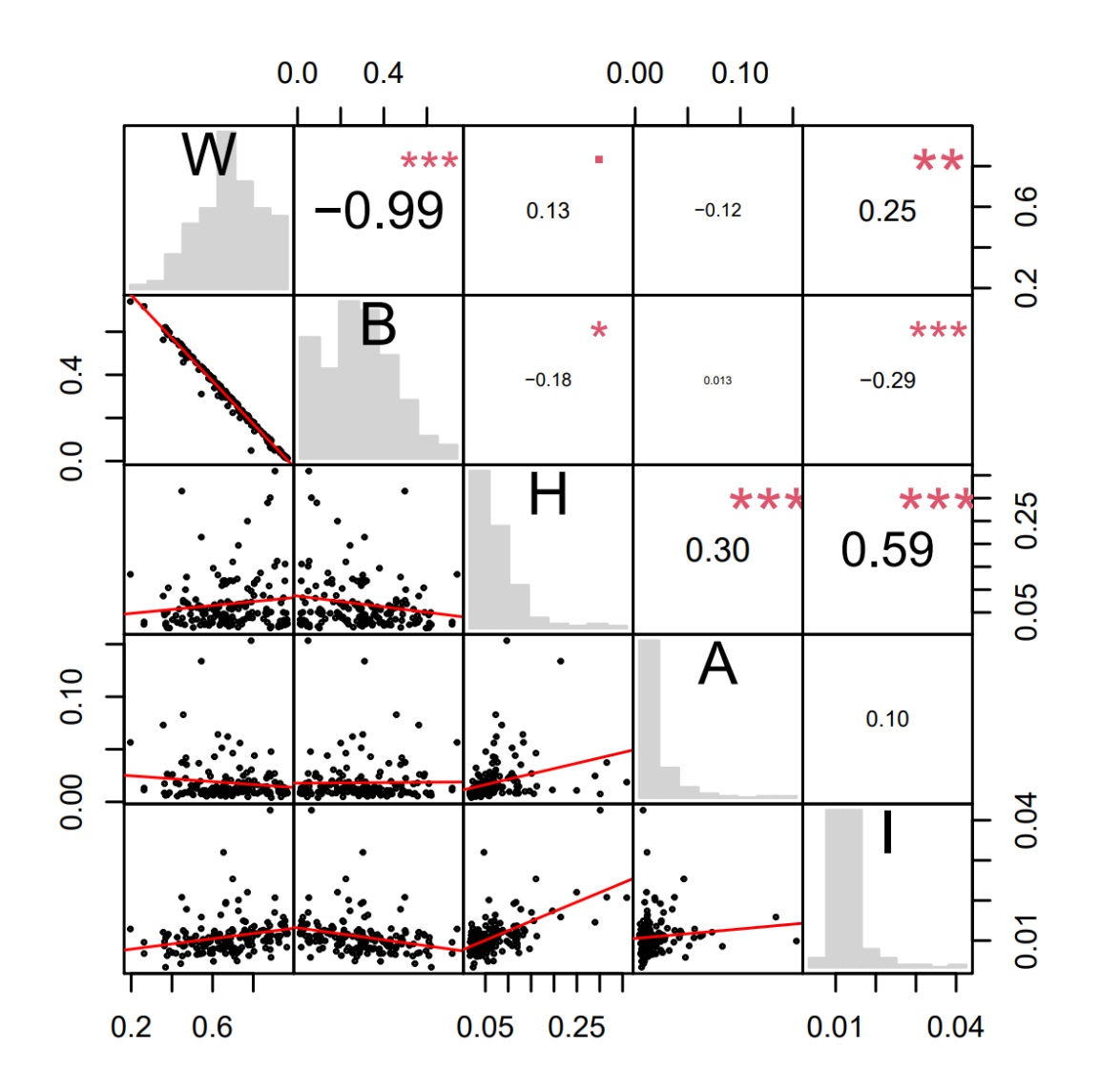

Supplemental Figure 1: Matrix of correlations between population proportions of census-reported race and ethnicity categories at the county level. Along the diagonal, histograms give the distribution of population proportions for the labeled racial and ethnic categories (W=White; B=Black or African American; H=Hispanic or Latino; A=Asian; I=American Indian or Alaska Native). Below the diagonal, scatterplots are given of pairs of these variables across counties, with the red line indicating the relationship determined by linear regression. Reflected over the diagonal, correlation coefficients are displayed with font size proportional to magnitude. Statistical significance is denoted using asterisks (.: *P* <0.10; *: *P* <0.05; **: *P* <0.01; ***: *P* <0.001).

**Table S1: Coefficients for demographic and epidemiological predictors of local nonpharmaceutical intervention public health orders.**

The best fit model for each response variable is given across a row, and the effect size for each predictor is given with a 95% confidence interval in parentheses. Statistical significance is denoted using asterisks (*: *P* <0.05; **: *P* <0.01; ***: *P* <0.001) and progressively darker shading corresponding to the same thresholds. Nagelkerke Pseudo-R^2^ values for each model are given in the second column. Predictors (from left to right) are: natural logarithm of population size (Population); median household income (Income); percent point difference of Republican and Democratic vote share in 2018 gubernatorial election (Partisanship); and date first case in county was detected (First Case).

|  | | **Demographic** | **Socioeconomic** | | **COVID-19** |
| --- | --- | --- | --- | --- | --- |
|  | **Pseudo-R^2^** | **Population** | **Income** | **Partisanship** | **First Case** |
| Social Distancing (all) | 0.31 | 0.56  (0.11, 1.06)* |  | -0.74  (-1.24, -0.26)** |  |
| Social Distancing (per capita deaths <2 per thousand) | 0.26 | 0.64  (0.17, 1.17)* |  | -0.68  (-1.20, -0.19)** |  |
| Social Distancing (per capita deaths <1 per thousand) | 0.25 | 1.01  (0.29, 1.85)* | -0.62  (-1.53, 0.11) | -0.66  (-1.24, -0.09)* |  |
| Shelter-in-Place (all) | 0.54 | 1.08  (0.23, 2.05)* | -0.55  (-1.29 , 0.07) |  | -1.90  (-3.19, -0.78)** |
| Shelter-in-Place (per capita deaths <1 per thousand) | 0.53 | 1.28  (0.36, 2.36)* | -0.46  (-1.18, 0.15) |  | -1.74  (-3.05, -0.59)** |
| Shelter-in-Place (per capita deaths <2 per thousand) | 0.51 | 1.29  (0.37, 2.37)* | -0.56  (-1.34, 0.08) |  | -1.84  (-3.19, -0.67)** |

**Table S2: Event study results for impact of social distancing and shelter-in-place orders on mobility.**

Fixed effects by policy day ($\beta_{p}$) with 95% confidence interval. Significance is denoted using asterisks (*: *P* <0.05; **: *P* <0.01; ***: *P* <0.001) and progressively darker shading corresponding to the same thresholds.

| **Policy Day** | **All Counties (Social Distancing)** | **All Counties (Shelter-in-Place)** |
| --- | --- | --- |
| 1 | -3.76 (-8.05, 0.53) | -3.03 (-7.05, 1.00) |
| 2 | -4.89 (-9.52, -0.24)* | -5.90 (-10.15, -1.65)** |
| 3 | -5.96 (-10.97, -0.94)* | -6.11 (-10.54, -1.68)** |
| 4 | -6.92 (-12.20, -1.64)* | -5.27 (-9.99, -0.55)* |
| 5 | -10.28 (-15.91, -4.65)*** | -3.61 (-8.52, 1.30) |
| 6 | -11.86 (-17.88, -5.84)*** | -0.97 (-6.06, 4.13) |
| 7 | -12.48 (-18.96, -6.00)*** | -4.08 (-9.30, 1.14) |
| 8 | -13.73 (-20.67, -6.78)*** | -5.88 (-11.53, -0.23)* |
| 9 | -14.97 (-22.62, -7.33)*** | -2.35 (-8.22, 3.53) |
| 10 | -18.86 (-27.23, -10.49)*** | -4.07 (-10.27, 2.13) |

**Table S3: Event study estimates of fixed effect of county on mobility.**

The names of all counties are given along with their corresponding estimates of fixed effect on mobility. Fixed effects were estimated separately for the model of the effects of social distancing and shelter-in-place and both values are given. Blank spaces indicate counties for which no mobility data were provided.

| **County** | **Social distancing** | **Shelter-in-Place** |
| --- | --- | --- |
| Appling | NA | NA |
| Atkinson | 10.13 | 10.9 |
| Bacon | 19.2 | 10.25 |
| Baker | NA | NA |
| Baldwin | 30.08 | 25.55 |
| Banks | 8.93 | 17.25 |
| Barrow | -16.27 | -10.55 |
| Bartow | -13.77 | -4.94 |
| Ben Hill | 2.93 | 8.5 |
| Berrien | -7.17 | -3.75 |
| Bibb | -8.57 | -8.4 |
| Bleckley | -18.87 | -17.2 |
| Brantley | -15.52 | -18.45 |
| Brooks | -16.57 | -12.15 |
| Bryan | -18.72 | -19.95 |
| Bulloch | 29.88 | 17.85 |
| Burke | 4.35 | -3.35 |
| Butts | -8.92 | -1.75 |
| Calhoun | NA | NA |
| Camden | -10.52 | -12.3 |
| Candler | -0.62 | 4.3 |
| Carroll | -15.07 | -9.41 |
| Catoosa | -10.87 | -6.2 |
| Charlton | -17.52 | -14.85 |
| Chatham | -10.18 | -19.7 |
| Chattahoochee | NA | NA |
| Chattooga | -17.22 | -6.75 |
| Cherokee | -33.67 | -28.11 |
| Clarke | 29.23 | 14.29 |
| Clay | NA | NA |
| Clayton | -26.01 | -34.41 |
| Clinch | -0.62 | 8.65 |
| Cobb | -30.33 | -32.06 |
| Coffee | 9.83 | 13.45 |
| Colquitt | -3.72 | 2.8 |
| Columbia | -10.92 | -9.95 |
| Cook | -5.87 | -3.8 |
| Coweta | -16.22 | -11.6 |
| Crawford | -3.37 | -7.75 |
| Crisp | 5.13 | 3.9 |
| Dade | -10.47 | -3.1 |
| Dawson | -14.87 | -9.15 |
| Decatur | 3.18 | 8.45 |
| Dekalb | -39.46 | -44.82 |
| Dodge | 1.58 | 4.8 |
| Dooly | 20.48 | 27.5 |
| Dougherty | -21.97 | -14.85 |
| Douglas | -28.12 | -22.48 |
| Early | -19.97 | -14.47 |
| Echols | 8.58 | 18.35 |
| Effingham | -11.62 | -10.5 |
| Elbert | 1.93 | 8.4 |
| Emanuel | -3.02 | -4.2 |
| Evans | 1.13 | 2.75 |
| Fannin | -2.22 | -3 |
| Fayette | -25.77 | -20.3 |
| Floyd | -2.32 | 3.18 |
| Forsyth | -28.92 | -24.5 |
| Franklin | 6.58 | 11.75 |
| Fulton | -25.4 | -36.02 |
| Gilmer | -3.67 | 1 |
| Glascock | NA | NA |
| Glynn | -4.37 | -4.2 |
| Gordon | -1.37 | 9.3 |
| Grady | -6.97 | -6.25 |
| Greene | -16.07 | -7.25 |
| Gwinnett | -31.53 | -33.34 |
| Habersham | -0.12 | 6.41 |
| Hall | -11.42 | -6 |
| Hancock | NA | NA |
| Haralson | 6.83 | 13.4 |
| Harris | 5.23 | 8.75 |
| Hart | -5.87 | -0.8 |
| Heard | 2.73 | 6.25 |
| Henry | -20.83 | -22.9 |
| Houston | -12.02 | -13.3 |
| Irwin | 13.68 | 14.9 |
| Jackson | -14.92 | -9 |
| Jasper | -9.77 | -5.55 |
| Jeff Davis | 4.13 | 7.1 |
| Jefferson | -4.12 | -5.6 |
| Jenkins | 1.08 | -4.05 |
| Johnson | -5.07 | 4.6 |
| Jones | -3.67 | 0.6 |
| Lamar | -16.87 | -13.75 |
| Lanier | -11.62 | -10 |
| Laurens | 2.73 | 5.65 |
| Lee | 6.33 | 15.45 |
| Liberty | -29.82 | -29.3 |
| Lincoln | -11.32 | -10.35 |
| Long | 16.68 | 20.2 |
| Lowndes | -6.22 | -0.72 |
| Lumpkin | 12.23 | 13.5 |
| Macon | -1.02 | -3.9 |
| Madison | -6.67 | 1.35 |
| Marion | NA | NA |
| Mcduffie | -1.72 | 3.4 |
| Mcintosh | 0.58 | -9.95 |
| Meriwether | -25.77 | -18.4 |
| Miller | -9.77 | -3.25 |
| Mitchell | -16.32 | -14.35 |
| Monroe | -5.02 | -1.3 |
| Montgomery | NA | 4.8 |
| Morgan | -12.47 | -5.35 |
| Murray | -3.67 | 1.9 |
| Muscogee | -18.32 | -19.5 |
| Newton | -21.62 | -16.6 |
| Oconee | -2.22 | 6.1 |
| Oglethorpe | -8.72 | -1.15 |
| Paulding | -22.82 | -15.3 |
| Peach | -5.97 | -5.15 |
| Pickens | -15.72 | -10.22 |
| Pierce | 4.98 | 7.25 |
| Pike | 5.03 | 10.4 |
| Polk | -20.12 | -11.39 |
| Pulaski | -22.12 | -16.4 |
| Putnam | -9.27 | -6.85 |
| Quitman | NA | NA |
| Rabun | -10.22 | -9.75 |
| Randolph | NA | NA |
| Richmond | -15.97 | -17.8 |
| Rockdale | -36.17 | -31.23 |
| Schley | NA | NA |
| Screven | -9.42 | -11.25 |
| Seminole | -5.52 | 5.15 |
| Spalding | -9.57 | -4.07 |
| Stephens | 0.78 | 5.3 |
| Stewart | NA | NA |
| Sumter | 10.18 | 10.35 |
| Talbot | -10.87 | -11.45 |
| Taliaferro | NA | NA |
| Tattnall | -20.87 | -16.9 |
| Taylor | -10.97 | -10.65 |
| Telfair | 4.68 | 5.95 |
| Terrell | -33.07 | -26.75 |
| Thomas | -0.97 | 2.7 |
| Tift | 8.33 | 11.98 |
| Toombs | 12.88 | 16 |
| Towns | 0.58 | -6.6 |
| Treutlen | -2.22 | 5 |
| Troup | 0.73 | 4.2 |
| Turner | 14.58 | 12.65 |
| Twiggs | NA | NA |
| Union | -1.12 | 0.05 |
| Upson | -4.92 | -0.6 |
| Walker | -10.02 | -5.05 |
| Walton | -14.32 | -8.25 |
| Ware | -0.02 | 3.4 |
| Warren | NA | NA |
| Washington | 4.78 | 8.8 |
| Wayne | 1.53 | 2.95 |
| Webster | NA | NA |
| Wheeler | NA | NA |
| White | -7.37 | -3.25 |
| Whitfield | -0.52 | 6.25 |
| Wilcox | -0.97 | -5.85 |
| Wilkes | -5.12 | -7 |
| Wilkinson | 12.33 | 13.85 |
| Worth | -24.77 | -20.25 |

**Table S4: Event study estimates of fixed effect of date on mobility.**

Fixed effects were estimated separately for the model of the effects of social distancing and shelter-in-place and both values are given. Blank spaces indicate dates that were not included in the given model (i.e., no county was within a ten-day time window of the public health order’s introduction).

| **Date** | **Social distancing** | **Shelter-in-Place** |
| --- | --- | --- |
| 3/1/2020 |  |  |
| 3/2/2020 |  |  |
| 3/3/2020 |  |  |
| 3/4/2020 |  |  |
| 3/5/2020 |  |  |
| 3/6/2020 |  |  |
| 3/7/2020 | 4.21 |  |
| 3/8/2020 | -41.14 | -28 |
| 3/9/2020 | -10.21 | -15 |
| 3/10/2020 | -11.31 | -10.66 |
| 3/11/2020 | -6.81 | -4.16 |
| 3/12/2020 | -6.98 | -3.66 |
| 3/13/2020 | -2.16 | 0.84 |
| 3/14/2020 | -36.36 | -56.14 |
| 3/15/2020 | -63.11 | -87.48 |
| 3/16/2020 | -37.2 | -62 |
| 3/17/2020 | -46.89 | -73.61 |
| 3/18/2020 | -51.41 | -78.91 |
| 3/19/2020 | -45.3 | -73.71 |
| 3/20/2020 | -42.01 | -66.12 |
| 3/21/2020 | -72.62 | -95.92 |
| 3/22/2020 | -96.53 | -121.89 |
| 3/23/2020 | -69.11 | -100.83 |
| 3/24/2020 | -68.69 | -97.48 |
| 3/25/2020 | -62.91 | -92.65 |
| 3/26/2020 | -61.57 | -92.36 |
| 3/27/2020 | -53.61 | -85.28 |
| 3/28/2020 | -72.52 | -107.31 |
| 3/29/2020 | -86.83 | -123.63 |
| 3/30/2020 | -61.49 | -98.92 |
| 3/31/2020 | -65.73 | -104.18 |
| 4/1/2020 | -56.07 | -96.38 |
| 4/2/2020 | -45.08 | -88.81 |
| 4/3/2020 |  | -94.38 |
| 4/4/2020 |  | -125.11 |
| 4/5/2020 |  | -133.8 |
| 4/6/2020 |  | -104.01 |
| 4/7/2020 |  | -105.55 |

**Table S5: Coefficients for socioeconomic predictors of mobility.**

The best fit model for each response variable is given across a row, and the effect size for each predictor is given with a 95% confidence interval. R^2^ values for each model are given in the second column. Significance is denoted using asterisks (*: *P* <0.05; **: *P* <0.01; ***: *P* <0.001) and progressively darker shading corresponding to the same thresholds. Predictors (from left to right) are aged-weighted infection fatality rates (Age); percent of population that is Black (Race); and median household income (Income).

|  | | **Demographic** | | **Socioeconomic** |
| --- | --- | --- | --- | --- |
|  | **R^2^** | **Age** | **Race** | **Income** |
| Final Mobility  (all counties) | 0.11 | -0.36 (-0.53, -0.20)*** | -0.22 (-0.39, -0.05)* | -0.22 (-0.39, -0.04)* |
| Final Mobility  (per capita deaths <2 per thousand) | 0.11 | -0.34 (-0.51, -0.17)*** | -0.19 (-0.37, -0.02)* | -0.24 (-0.39, -0.03)* |
| Final Mobility  (per capita deaths <1 per thousand) | 0.13 | -0.34 (-0.51, -0.17)*** | -0.17 (-0.36, 0.01) | -0.23 (-0.41, -0.04)* |

**Table S6: Coefficients for socioeconomic, health, and legislative predictors of early epidemiological outcomes.**

The best fit model for each response variable is given across a row, and the effect size for each predictor is given with a 95% confidence interval. Significance is denoted using asterisks (*: *P* <0.05; **: *P* <0.01; ***: *P* <0.001) and progressively darker shading corresponding to the same thresholds. Nagelkerke Pseudo-R^2^ values for each model are given in the second column in bold. Predictors (from left to right) are aged-weighted infection fatality rates (Age); percent of population that is Black (Race); natural logarithm of population size (Pop.); age-adjusted emergency room visit rate for asthma (Asthma); prevalence of diabetes in adults (Diab.); coronary heart disease-related hospitalization rate (C.H.D.); annual average ambient PM2.5 concentration (Poll.); percent of population with a high school degree (Edu.); proportion of population living in poverty (Poverty); unemployment rate (Unemp.); proportion of population living in poverty (Poverty); average normalized daily mobility in the final week of April (Mob.); date first case in county was detected (F.C .).

|  | | **Demographic** | | | **Health** | | | **Socioeconomic** | | | | **Behav-**  **ior** | **COVID-19** |
| --- | --- | --- | --- | --- | --- | --- | --- | --- | --- | --- | --- | --- | --- |
|  | **Pseudo-R^2^** | **Age** | **Race** | **Pop.** | **Asthma** | **Diab.** | **C.H.D.** | **Poll.** | **Edu.** | **Poverty** | **Unemp.** | **Mob.** | **F.C** |
| Cases (all counties) | 0.73 | 0.16  (-0.01,  0.33)  * |  | 0.81  (0.62, 1.00)  *** |  |  | -0.23  (-0.35,  -0.11)  *** | 0.24  (0.10,  0.38)  *** |  | 0.37 (0.24, 0.50)  *** |  |  | -0.22  (-0.39,  -0.05) ** |
| Cases (per capita deaths <2 per thousand) | 0.68 | 0.16  (-0.01,  0.32)  * |  | 1.00  (0.83, 1.18)  *** |  | 0.10  (-0.01, 0.22) | -0.20  (-0.32,  -0.08)  ** | 0.25  (0.11, 0.38)  *** |  | 0.32  (0.20, 0.45)  *** |  |  |  |
| Cases (per capita deaths <1 per thousand) | 0.67 | 0.23  (0.85,  0.37)  ** | 0.20  (0.04, 0.35)  ** | 0.89  (0.75, 1.04)  *** |  | 0.14 (0.03, 0.25)  * |  | 0.16  (0.03, 0.30)  ** |  |  | -0.13  (-0.28, 0.03) | 0.11  (0.00, 0.22)  * |  |
| Deaths (all counties) | 0.53 |  |  | 0.62  (0.34,  0.91)  *** | 0.17  (-0.05,  0.40) |  | -0.25  (-0.42,  -0.07)  * | 0.27  (0.06,  0.48)  ** | -0.23  (-0.50,  0.03) | 0.32  (0.05,  0.60)  * |  |  | -0.43  (-0.71,  -0.18)  *** |
| Deaths (per capita deaths <2 per thousand) | 0.48 |  |  | 0.75  (0.45,  1.05)  *** |  |  | -0.23  (-0.41,  -0.05)  * | 0.30  (0.09,  0.51)  ** | -0.22  (-0.49,  0.05) | 0.40  (0.17,  0.63)  *** |  |  | -0.34  (-0.63,  -0.05)  * |
| Deaths (per capita deaths <1 per thousand) | 0.49 |  |  | 0.72  (0.46,  0.99)  *** | 0.24  (0.04, 0.44)  * |  |  | 0.29  (0.09,  0.49)  ** | -0.24  (-0.46,  -0.03)  * |  | -0.18  (-0.43,  0.06) | 0.19  (0.02,  0.37)  * | -0.20  (-0.46, 0.06) |

**Table S7: Data dictionary. Names of all predictors and response variables referenced in the texts, along with detailed descriptions of the variable.**

| **Name** | **Description** |
| --- | --- |
| Age | aged-weighted infection fatality rates; predictor |
| Race | percent of population that is Black; predictor |
| Population (Pop.) | natural log of population size; predictor |
| Asthma | age-adjusted emergency room visit rate for asthma per 100,000 people; predictor |
| Diabetes (Diab.) | prevalence of diabetes in adults; predictor |
| Coronary Heart Disease (C.H.D.) | Coronary heart disease-related hospitalization rate per 1,000 Medicare beneficiaries over age 65+; predictor |
| Pollution (Poll.) | annual average ambient PM2.5 concentration; predictor |
| Education (Edu.) | percent of population with a high school degree; predictor |
| Income | median household income (thousands of dollars); predictor |
| Partisanship | percent point difference of Republican and Democratic vote share in 2018 gubernatorial election; continuous predictor variable; predictor |
| Poverty | Percent of population living in poverty; predictor |
| Unemployment  (Unemp.) | unemployment rate; predictor |
| First Case (F.C.) | date first case in county was detected; predictor |
| Social Distancing | binary whether policies were introduced at either the county level prior to the statewide order to encourage social distancing in the general population (e.g., ban on gatherings at non-essential businesses, restrictions on gathering sizes, closure of public use areas); binary response variable for analysis 1 |
| Shelter-in-Place | whether a county level shelter-in-place was introduced at the county level prior to the statewide order; binary response variable for analysis 1 |
| Mobility (Mob.) | average daily mobility (defined as the median radius of movement across devices in a county) in the final week of April as a proportion of a pre-pandemic baseline between February 17^th^ and March 17^th^, 2020; continuous response variable for analysis 3 and predictor for analysis 4 |
| Cases | Cumulative COVID-19 cases reported within four weeks of a county’s first detected case; discrete response variable for analysis 4 |
| Deaths | Cumulative COVID-19 deaths reported within six weeks of a county’s first detected case; discrete response variable for analysis 4 |

1	Gray DM, Anyane-Yeboa A, Balzora S, Issaka RB, May FP. COVID-19 and the other pandemic: populations made vulnerable by systemic inequity. *Nature Reviews Gastroenterology & Hepatology* 2020: 1–3.

2	Williams DR, Cooper LA. COVID-19 and health equity—a new kind of “herd immunity”. *JAMA* 2020; **323**: 2478–80.

3	van Holm EJ, Wyczalkowski CK, Dantzler PA. Neighborhood conditions and the initial outbreak of COVID-19: the case of Louisiana. *Journal of Public Health* 2020; : 1–6.

4	Azar KMJ, Shen Z, Romanelli RJ, *et al.* Disparities in outcomes among COVID-19 patients in a large health care system in California. *Health Affairs* 2020; **39**: 1253–62.
